# Potential public health and economic impact of influenza and COVID-19 multicomponent vaccination (mRNA-1083) for adults aged 65 years and older in France

**DOI:** 10.64898/2026.11.25.26368982

**Authors:** D Mehta, L Biron, J Carette, C Demont, G Gavazzi, O Guerin, H Leleu, O Reynaud Levy, O Rozaire, M Uhart, N Van de Velde, E Beck

**Affiliations:** Moderna Inc., Cambridge MA, USA; Cencora, Paris, France; Moderna Inc., Paris, France; CHU Grenoble-Alpes, Grenoble, France; CHU Nice, Nice, France; Assistance Publique des Hôpitaux de Marseille, Marseille, France; Union Régionale des Professionnels de Santé Pharmaciens, Auvergne-Rhône Alpes, Lyon, France

**Keywords:** COVID-19, influenza, France, mRNA-1083, public health, costs, vaccination

## Abstract

Seasonal influenza and SARS-CoV-2 continue to cause substantial disease burden, particularly among older adults. The potential public health and economic impact of introducing combined enhanced influenza/COVID-19 vaccine, mRNA-1083, was assessed, for adults ≥65 years, versus separate high-dose inactivated influenza vaccine (HD-IIV) and BNT162b2 COVID-19 vaccination. A static decision-tree health economic model for France evaluated five value dimensions with mRNA-1083, in a stepwise approach, during an average post-pandemic season. Health system efficiencies, potential improved protection versus current comparators, avoidance of influenza egg-adaptation effects, and increased vaccine coverage rates (VCR) were assessed in the base case, and a future-looking scenario assessed potential reduced strain mismatch from antigenic drift, due to later influenza strain selection enabled by the mRNA platform. During an average season, mRNA-1083 was estimated to potentially prevent 25,830 medically-attended cases and 4,286 hospitalisations, saving €69.4 million in healthcare costs. Replacing separate influenza and COVID-19 vaccinations with mRNA-1083 may generate health system efficiencies, including 439,285 healthcare-provider hours saved. Potential improved clinical protection with mRNA-1083 prevented 2,194 hospitalisations saving €11.8 million in treatment costs. Avoiding egg-adaptation effects prevented a further 75 influenza hospitalisations, saving €0.3 million. Increased VCR with mRNA-1083 prevented an additional 2,017 hospitalisations, saving €10.4 million. Later strain selection could reduce antigenic drift and prevent a further 148 influenza hospitalisations and €0.6 million treatment cost on average per season. Results support that mRNA-1083 may improve seasonal respiratory vaccination by combining convenience, health system efficiency and potential for improved protection, higher coverage, and better influenza strain matching in a single injection.

## Introduction

Seasonal influenza virus and SARS-CoV-2 cause widespread respiratory infections with recurring seasonal patterns [1–4], and older adults bear a disproportionate burden, with higher risks of severe disease, hospitalisation, and death [5–7]. The 15 million (M) adults currently aged ≥65 (22% of the population) and the ageing population in France pose a challenge for the healthcare system, especially in winter, with co-circulation of respiratory viruses [5, 8].

Seasonal influenza affects 2–6M individuals annually in France [5]. The 2024/25 season was more severe than previous years, with early onset, rapid case increases, and co-circulation of three influenza subtypes; A(H1N1), B Victoria, and A(H3N2), each accounting for around a third of cases [9]. During the season, influenza resulted in approximately 2.7M primary care visits and 224,000 emergency visits for influenza-like illness (ILI), alongside over 29,100 hospitalisations following emergency visits, 60% among ages ≥65 years [9–11]. The national hospital discharge database (Programme de Médicalisation des Systèmes d’Information, PMSI) reported 36,440 influenza hospitalisations in 2024 among adults ≥65 years [12]. Overall, there were around 9,000 influenza deaths (92% in adults ≥65 [10]) and an estimated 17,600 excess deaths (all-cause) during the 2024/25 influenza epidemic season (compared with 8,100 during 2023/24) [11]. COVID-19 is now endemic in Europe, and appears to have continuous year-round circulation with larger winter peaks as well as summer peaks [13]. While there are fewer cases than during the pandemic, at-risk populations still experience an important burden; with an estimated 287,289 primary care attended cases [9] and with 107,742 hospitalisations in France among older adults reported in PMSI in 2024 [12]. Santé Publique France estimated that COVID-19 was responsible for 2,887 deaths (97% in adults ≥65 [6]). Surveillance data from 2023/24 showed older adults accounted for 63% of intensive care admissions [14].

Vaccination plays a critical preventive role in reducing influenza and COVID-19 burden in France, particularly among older adults. However, coverage rates remain below targets (75% for influenza [15]), with only 53.7% vaccinated against influenza and 21.7% against COVID-19 among adults ≥65 years in the 2024/25 season [16]. Current strategies prioritise reducing severe outcomes in high-risk populations [17], recommending enhanced (e.g., high-dose [HD]) inactivated influenza vaccines (IIV) in older adults [18] and COVID-19 vaccination with mRNA vaccines, mRNA-1273 (Spikevax, Moderna) and BNT162b2 (Comirnaty, Pfizer/BioNTech), or protein-based vaccine NVX-CoV2705 (Nuvaxovid, Sanofi) [19, 20]. Of note, co-vaccination of influenza and COVID-19 vaccines is also recommended in France, as a strategy to simplify vaccine administration [21].

A combined influenza/COVID-19 vaccine could improve vaccination coverage by reducing the number of injections and simplifying delivery. Preference studies in the United States [22] and Europe (including in France) [23] showed high acceptability among adults ≥65 years and healthcare professionals (HCPs), driven by convenience and dual protection. Findings suggested potential increases in coverage of around 2–12 percentage points for COVID-19 and 3–10 points for influenza [23], supported by positive perceptions from French pharmacists gathered from market research regarding improved compliance and efficiency [24].

mRNA-1083 (mCombriax, Moderna) is the first combination vaccine, delivered in a prefilled syringe, indicated for active immunisation against influenza and COVID-19 in adults aged ≥50 years [25]. It combines an enhanced influenza component (mRNA-1010, targeting A/H1N1, A/H3N2 and B/Victoria [26, 27]); and a COVID-19 component designed to target immunodominant epitopes (mRNA-1283, mNexspike, Moderna [28]).

Beyond the operational advantages of a combination vaccine, the mRNA platform offers additional benefits for influenza vaccination. Conventional influenza vaccines are predominantly produced using egg-based manufacturing processes, which can introduce egg-adaptive mutations that alter viral antigens and reduce the match between vaccine strains and circulating viruses [29]. By contrast, mRNA influenza vaccines are cell-free and avoid egg adaptation. Moreover, the rapid manufacturing process enabled by mRNA technology allows for later strain selection than in February and, thus, based on more recent global surveillance data, has the potential to improve strain matching and vaccine effectiveness in seasons characterised by antigenic drift [30].

In a pivotal phase 3 randomised controlled trial (NCT06097273), mRNA-1083 induced stronger immune responses against all three clinically-relevant and WHO-recommended influenza strains (A/H1N1, A/HN2, and B/Victoria) compared with the vaccine recommended in France for adults ≥65 years (HD-IIV, Efluelda, Sanofi); and against mRNA-1273 vaccine for COVID-19 [31]. The influenza antigen design in the mRNA component of mRNA-1083 is the same antigen design as in mRNA-1010. mRNA-1010 demonstrated superior vaccine efficacy (relative vaccine efficacy [rVE] of 26.6%) versus SD-IIV in adults 50 years [32]; and superior immunogenicity to HD-IIV in adults aged ≥65 years [26] in pivotal phase 3 trials, and was recently recommended by the American Academy of Family Physicians as an enhanced influenza vaccine [27]. Similarly for COVID-19, mRNA-1083 uses the same antigen design as mRNA-1283. mRNA-1283 showed non-inferiority, higher immune responses, and numerically higher rVE (13.5%, 95% CI -7·7–30·6) versus mRNA-1273 in adults ≥65 years in a pivotal phase 3 trial (NextCOVE [28]). A recent real-world evidence (RWE) study (2025/2026 season) among US Veterans reported an rVE of 35.9% (against medically-attended COVID-19) and 47.0% (against COVID-19 hospitalisations) for mRNA-1283 compared with BNT162b2 [33].

mRNA-1083 has the potential to provide additional value through three complementary mechanisms: (i) improving healthcare system efficiency by replacing two separate vaccinations with a single injection; (ii) providing improved clinical protection through enhanced influenza and immunodominant epitope targeting COVID-19 vaccine components; and better strain match by preventing egg-adaptation and allowing later strain selection; and (iii) increasing seasonal vaccination coverage by simplifying vaccine delivery and improving convenience for vaccine recipients and HCPs. These mechanisms have the potential to generate incremental public health and economic benefits beyond those achievable with separate influenza and COVID-19 vaccination programs.

The objective of this study was to assess the potential public health and economic impact of introducing combination influenza and COVID-19 vaccination with mRNA-1083, for adults

≥65 years in France in an average post-pandemic season (October to September), compared with recommended vaccines for influenza (HD-IIV) [34] and for COVID-19 (BNT162b2).

## Methods

### Model description

A static decision-tree health economic model [35–37] was adapted to compare vaccination with mRNA-1083 versus HD-IIV and BNT162b2, in the French population of adults aged ≥65 years (N=14,924,665 [38]). The impact of vaccination was projected over an average season (October 1^st^ to September 30^th^) informed by medically-attended infection rates in the post-pandemic setting.

The model includes a probability of developing symptomatic influenza or COVID-19 infections requiring either no formal care, outpatient care, or hospitalization (with or without intensive care unit [ICU] or ventilator, and potential in-hospital death), followed by a probability of readmission, post-COVID condition or post-discharge death. The likelihood of developing an infection and of being hospitalized following infection was reduced in vaccinated individuals and varied with each vaccine (**Figure 1**).

**Figure 1.**
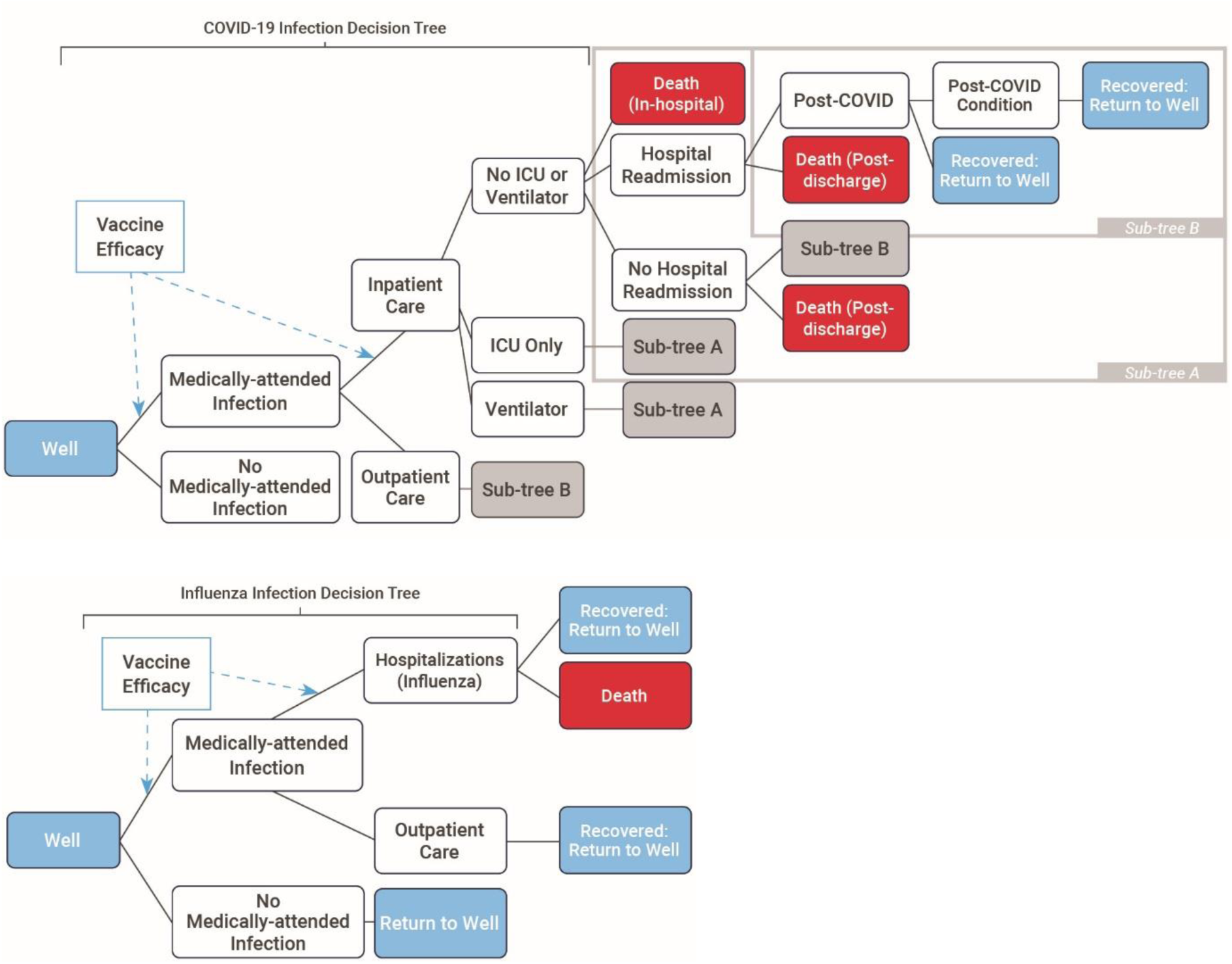
COVID-19 and influenza model structures (based on [35]) Note: Outpatient care includes physician visits and emergency department visits; ICU: intensive care unit

### Model inputs and assumptions

To account for the ongoing variability in COVID-19 epidemiology, the model represented an average post-pandemic season using the mean medically-attended COVID-19 infection rate observed during the 2023/24 and 2024/25 seasons in France [39], consistent with the approach used for seasonal influenza modelling. The model also considered the 2024/25 COVID-19 vaccination coverage rate (VCR) of 21.7% [16] and real-world vaccine effectiveness (VE) for mRNA-1273 vaccination in 2024/25 [40] to estimate the rate of medically-attended COVID-19 infections with no vaccination, stratified by month. A weighted average, based on the seasonality observed each year, was also used to estimate the annual number of COVID-19 related events stratified by level of care (i.e., hospitalizations, ICU admissions, outpatient visits, and deaths) [9, 41–43]. The risks of post-covid condition, including long COVID [44] and myocarditis due to COVID-19 infections [45], were also included. Similarly for influenza, the average of 2023/24 and 2024/25 influenza epidemic seasons was used for medically-attended infections and related events [9, 42, 43], with the rate of medically-attended influenza infections with no vaccination estimated considering annual influenza VCR in 2024/2025 (53.7% [16]) and the VE of standard-dose vaccines (22% [46]), as no high-dose influenza vaccines were available yet [11]. This model calibration was performed for adults aged 65–74 years and ≥75 years (**Table S1**).

Health benefits of vaccination were presented as quality-adjusted life-year (QALY) gains, using baseline utility values from the French general population [47], and published utility loss values (from other countries) associated with symptomatic influenza and COVID-19 infections [48, 49], influenza hospitalization [50], COVID-19 hospitalization (with or without ICU admission) [51], long COVID after outpatient care or hospitalization [52], myocarditis due to COVID-19 disease [53].

For the public health and economic impact analysis, the model compared the average vaccine administration costs (from administration by a pharmacist, nurse or doctor [54–56]) and COVID-19 and seasonal influenza treatment costs, due to outpatient care (GP consultation and paracetamol [56, 57]), hospitalizations [58, 59], and long COVID care [60], with and without mRNA-1083 vaccination (**Table S1**). All costs were inflated to 2025 Euro values, using the Consumer Price Index [61].

### Vaccination strategy and parameters

The analysis adopted a stepwise approach to evaluating the incremental value associated with the mRNA-1083 introduction. The potential benefits considered in the base case included greater convenience for vaccine recipients and improved efficiency for HCPs and health systems, clinical benefits relative to current influenza and COVID-19 vaccine comparators, mRNA-platform advantages preventing egg-based adaptations, and the potential for increased COVID-19 and seasonal influenza vaccine coverage. An additional scenario assessed mRNA-1083 prevention of strain mismatch due to the potential for later strain selection enabled by the mRNA platform. Specifically, the five potential value dimensions explored in the model in sequential steps were (1) system efficiencies arising from administration of a single combination vaccine, (2) the clinical benefit associated with the potential more robust protection provided by mRNA-1083 compared with current influenza and COVID-19 vaccine comparators, (3) the absence of egg-adaptation, (4) the potential impact of increased seasonal vaccination coverage resulting from the convenience of a combination vaccine; and (5) as an additional scenario, the potential benefit associated with late strain selection. Assessing these components stepwise enabled the contribution of each parameter to the overall public health and economic outcomes to be quantified independently, as well as evaluating their overall effect.

#### Value dimension 1: Health system efficiencies

The operational efficiencies of replacing separate influenza and COVID-19 vaccinations with a single mRNA-1083 injection were quantified; the combination vaccine was assumed to eliminate one vaccine administration per vaccinated individual.

Health system efficiencies were modelled by incorporating reductions in HCP time and vaccine administration costs. Based on published evidence, administration of mRNA-1083 was assumed to save up to 11 minutes of HCP time compared with the delivery of separate influenza and COVID-19 vaccines [62]. In addition, the cost of one vaccine administration was avoided for each individual receiving mRNA-1083.

#### Value dimension 2: More robust protection

The potential clinical benefit associated with the increased protection conferred by mRNA-1083 compared with currently recommended enhanced influenza and COVID-19 vaccines was evaluated. Vaccine effectiveness (VE) inputs for mRNA-1083 were informed by evidence from pivotal phase 3 clinical trials, correlate-of-protection (CoP) analyses, and real-world evidence (RWE). In pivotal phase 3 trials, mRNA-1083, and mRNA-1010 and mRNA-1283 which share the same encoded antigens as mRNA-1083, demonstrated superior immunogenicity versus HD-IIV in adults aged ≥65 years for clinically-relevant influenza strains, and higher immunogenicity versus mRNA-1273 in individuals aged ≥65 years, respectively [26, 28, 31]. For influenza, a correlate-of-protection (CoP) analysis estimated the rVE of mRNA-1010 versus HD-IIV as 12.4% (95% CI 3.4–21.3%) against any medically-attended influenza in adults aged ≥65 years [63]. For COVID-19, a retrospective matched cohort study among US veterans aged ≥65 years during 2025/2026 season estimated that the comparative effectiveness of mRNA-1283 versus BNT162b2 was 35.9% (95% CI 20.4–49.7) against medically-attended COVID-19 and 47.0% (95% CI 5.4–74.8) against COVID-19 hospitalization [33].

In this analysis, mRNA-1083 was compared to HD-IIV and BNT162b2, used as proxies for enhanced seasonal influenza and mRNA COVID-19 vaccines, respectively. The model VE of mRNA-1083 against seasonal influenza was estimated by applying the mRNA-1010 versus HD-IIV rVE CoP estimate of 12.4% (95% CI 3.4–21.3%) [63] to the initial RWE based VE for HD-IIV [64]. The VE of mRNA-1083 against COVID-19 was based on the RWE (US 2025/2026 season) VE of mRNA-1283 against COVID-19 hospitalization 59.3% (95% CI 39.0–72.9%) and COVID-19 medically-attended infections 42.0% (95% CI 35.0–48.3%) in adults ≥ 65 years [65]. Next, to derive the VE of BNT162b2, mRNA-1283 versus BNT162b2 rVE RWE estimates [33] were applied to these initial RWE based COVID-19 VE values [65] (**Table 1)**.

**Table 1.** rVE of mRNA-1083 versus BNT162b2 and HD-IIV.

| **Average VE** | **Initial VE (Month 1)** | **rVE mRNA-1083** |
|---|---|---|
| **Influenza vaccination** |  |  |
| **mRNA-1083** |  |  |
| Infection: 46.2% (7m) | 70.2% (CoP rVE) |  |
|  | 72.7% (CoP rVE+ egg-based rVE) |  |
| Hospitalization: 46.2% (7m) | 70.2% (CoP rVE) |  |
|  | 72.7% (CoP rVE+ egg-based rVE) |  |
| **HD-IIV** |  |  |
| Infection: 42.0% [64] | 66.0% (Monthly waning 8% over 7m) | 12.4% CoP [68]<br>8.3% egg-adaptation [29, 30, 37] |
| Hospitalization: 42.0% [64] | 66.0% (Monthly waning 8% over 7m) |  |
| **COVID-19 vaccination** |  |  |
| **mRNA-1083** |  |  |
| Infection: 25.6% (12m) | 51.7% [65] |  |
| Hospitalization: 50.8% (12m) | 64.3% [65] |  |
| **BNT162b2** |  |  |
| Infection: 6.4% (12m) | 24.6% | 35.9% [33] |
| Hospitalization: 19.2% (12m) | 32.7% | 47.0% [33] |

m: months; (r)VE: (relative) vaccine effectiveness

The model considered an initial VE and a monthly waning of protection for influenza and COVID-19 vaccines. Thus, HD-IIV initial VE was estimated at 66.0% for both seasonal influenza hospitalization and medically-attended infections, based on the observed average VE of HD-IIV (42.0% [64]) over 7 months and a monthly waning rate of 8% [66, 67]. The initial VE of mRNA-1083 [65] was estimated at 64.3% and 51.7% against COVID-19 hospitalizations and medically-attended infections, considering a follow-up of 69 days and a monthly waning assumed at 2.46% for hospitalizations and 4.75% for infections [65]. rVEs were applied during the initial month, following previously described methods [36].

#### Value dimension 3: Absence of egg adaptation

Additional influenza-related value may arise from the potential increase in influenza VE associated with the non-egg-based manufacturing process of mRNA-1083. Unlike egg-based influenza vaccines such as HD-IIV, mRNA-1083 is not subject to egg-adaptive mutations that may reduce antigenic match between vaccine and circulating strains leading to a reduction in VE. A review reported evidence of egg adaptation affecting VE in 6 of 9 (67%) Northern Hemisphere influenza seasons between 2011/12 and 2019/20 [30]. Across seasons in which egg adaptation was observed, the rVE associated with avoiding egg adaptation was estimated at 12.4% [29]. To represent an average influenza season, this rVE was weighted by the probability of egg adaptation occurring in a given season [30], resulting in an average rVE of 8.3% [37]. This rVE was then applied to the VE of mRNA-1083, as the CoP-derived rVE from step 2 was based on clinical data during seasons in which no egg-adaptation was observed (Table 1).

#### Value dimension 4: Improving Vaccination Coverage Rates (VCR)

mRNA-1083 may also increase public health impact by improving vaccine coverage rates (VCR). Based on the preference study results suggesting the majority of the French surveyed population would be willing to get a combination vaccine [23], a 10 percentage-point increase in COVID-19 vaccine coverage was assumed, from 21.7% to 31.7%, and a 5 percentage-point increase in influenza vaccine coverage, from 53.7% to 58.7%. The resulting improved VCR for mRNA-1083 was therefore 31.7%, with the remaining influenza coverage assigned to HD-IIV, corresponding to 27.0% of the population (58.7% minus 31.7%, **Figure S1**).

#### Value dimension 5 (scenario): Genetic Drift and Late Strain Selection

Each year, the World Health Organization (WHO) provides recommendations in February or March for separate influenza strains for egg-based and recombinant-based (RIV3) vaccines, to allow sufficient time for vaccine manufacturing prior to the start of the vaccination season in September. The strains of the three viral types (A/H1N1, A/H3N2, B-Victoria) included in the seasonal influenza vaccines are reassessed annually as influenza viruses constantly mutate. During the six-month lag between March and September, circulating viruses may become antigenically different or “mismatched” (genetic drift) from the WHO and regulators’ selected vaccine viruses. A/H3N2, typically the dominant circulating influenza strain, mutates more quickly than A/H1N1 or B/Victoria and is therefore most prone to genetic drift [69]. A review of circulating strains in nine previous influenza seasons (2011/12 to 2019/20) found that in two seasons (2014/15 and 2018/19) A/H3N2 drifted early enough in the season to impact VE [30]. As mRNA-1010 can be manufactured more quickly than other influenza vaccines, the choice of virus for these vaccines may be postponed by three months, until end of May, allowing for a reduced probability of mismatch between vaccine and circulating strains. In the model, the likelihood of a genetic drift occurring in a season was based on the average of two in nine seasons [30], assuming strain mismatch with HD-IIV vaccination in seasons with a genetic drift but not with mRNA-1083, due to the ability for rapid strain adaptation and manufacturing[36]. The potential difference in VE with mRNA-1083 versus HD-IIV during a strain mismatch season was based on the French estimate of 15 percentage points [70]. Thus, for the projected season an average difference in VE of 3.3% was modelled. As implementation of later strain selection for seasonal influenza vaccines would require changes to the current WHO strain selection timeline and associated regulatory and policy processes, this benefit is likely to be achieved in the middle-to long-term. Consequently, the potential impact of reduced strain mismatch was only considered as a future-looking scenario analysis.

### Scenario analyses

In the base case analysis considering steps 1–4, the introduction of mRNA-1083 was assumed to increase vaccination coverage rates (VCRs) for both COVID-19 and influenza through improved vaccine uptake. To explore the impact of this assumption, the VCR was varied in scenario analyses. A conservative ‘COVID-19 VCR’ scenario assumed mRNA-1083 would have no impact on current COVID-19 and influenza VCRs; thus, the same VCR was assumed (21.7%) for mRNA-1083 and BNT162b2, and the remaining 32.0% was assumed for HD-IIV to match the observed influenza VCR, i.e. 53.7%-21.7%=32.0%. An ‘Influenza VCR’ scenario was also assessed, assuming mRNA-1083 VCR would match current influenza VCR (53.7%), resulting in a significant increase in COVID-19 VCR and no change to influenza VCR (**Figure S1**).

Additional scenario analyses assessed the impact of varying assumptions on: COVID-19 and seasonal influenza medically-attended infection rates (by +/-25%); varying rVEs using their 95% CI, for seasonal influenza CoP rVE 12.4% (95% CI 3.4–21.3) against any medically attended seasonal influenza, COVID-19 rVE 35.9% (95% CI 20.4–49.7) against medically-attended COVID-19 and 47.0% (95% CI 5.4–74.8) against hospitalization; and varying the egg-adapted rVE between 4.1% and 12.4%.

## Results

mRNA-1083 vaccination was estimated to provide considerable public health and economic benefits versus separate HD-IIV and BNT162b2 vaccinations, during an average seasonal influenza/COVID-19 season in France. In the base case (steps 1–4), mRNA-1083 prevented an additional 25,830 medically attended infections, 4,286 hospitalizations and 1,331 deaths in an average season, with a total 7,680 QALYs prevented and healthcare cost savings of €69,362,502, including administration and treatment cost savings (**Figure 2**).

**Figure 2.**
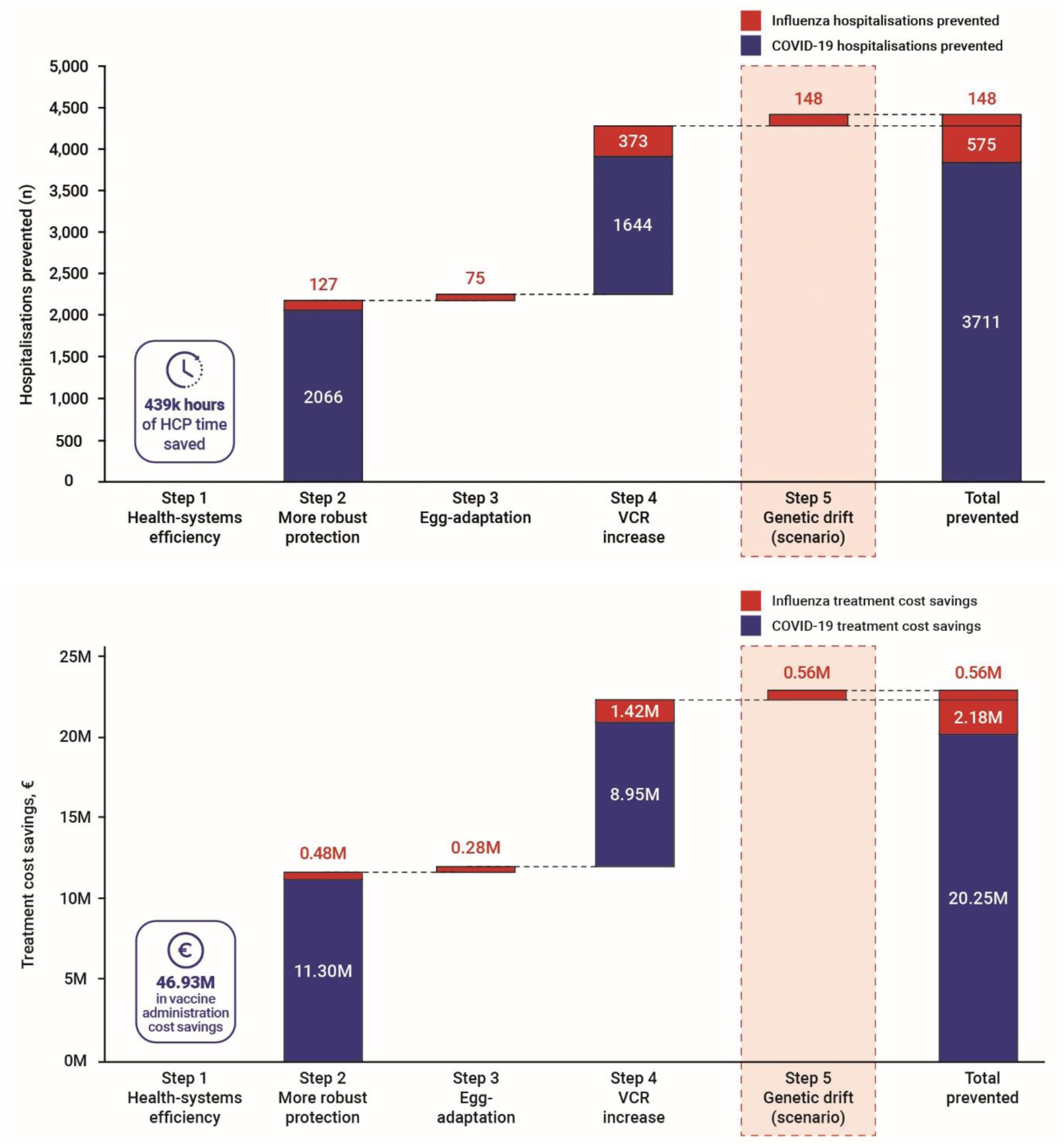
a) Hospitalizations prevented and b) treatment cost savings for mRNA-1083 versus BNT162b2 and HD-IIV in five incremental steps. Note: Total COVID-19 hospitalizations prevented (3710.8) was rounded up to 3711 above; HCP: healthcare provider; k: thousand; M: million; n: number; VCR: vaccine coverage rate; Impact of vaccine in egg-adaptation and genetic drift scenario assumed for average season

The contribution of each value dimension to these outcomes was evaluated sequentially. Health system efficiencies were estimated to save 439,285 hours of HCP time by converting the 16.1% of adults who received both influenza and COVID-19 vaccinations (based on 2024/2025 season VCR data) to a single mRNA-1083 administration, thus halving the overall number of vaccine administrations. Vaccine administration cost-savings were €60,983,823.

Enhanced clinical protection, assuming a VCR of 21.7% for mRNA-1083 (matching current COVID-19 VCR) and 32.0% for HD-IIV (to match current total influenza VCR), resulted in 2,194 fewer hospitalizations (127 and 2,066 for influenza and COVID-19, respectively). Corresponding treatment cost-savings amounted to €11,296,805 for COVID-19 and €483,569 for influenza.

Considering the impact across multiple seasons, avoidance of egg adaptation was estimated to prevent on average per season a further 75 influenza-related hospitalizations compared with HD-IIV, generating additional influenza treatment cost savings of €283,265.

Increased vaccination coverage (10 percentage-point increase in COVID-19 VCR to 31.7% and 5 percentage-point increase in influenza VCR to 58.7%), informed by preference study estimates, resulted in a further 2,017 hospitalizations avoided (373 influenza and 1,644 COVID-19). Additional treatment cost savings were estimated at €8,949,172 for COVID-19 and €1,417,418 for influenza.

Finally, a future-looking scenario on later strain selection enabled by the rapid manufacturing process of the mRNA platform was estimated to prevent on average per season an additional 148 influenza-related hospitalizations by reducing the impact of antigenic drift. This translated into further influenza treatment cost savings of €564,068.

Results of symptomatic infections and deaths prevented, and quality-adjusted life-years (QALYs) saved, with mRNA-1083 compared with BNT162b2 and HD-IIV by step are presented in **Table S2**.

The results of the scenario analyses are presented as incremental hospitalizations prevented and treatment cost savings relative to the base case, which incorporated value dimensions 1–4 (**Figure 3**). Across all scenarios, mRNA-1083 remained associated with additional health benefits and healthcare cost savings compared with separate HD-IIV and BNT162b2 vaccination. The results were most sensitive to assumptions regarding VCR, COVID-19 rVE, and COVID-19 incidence. The greatest benefits were observed when mRNA-1083 VCR was assumed to match the current influenza VCR (53.7%), resulting in 7,828 hospitalizations prevented and approximately €42 million in treatment cost savings. Even when assuming no increase in VCR compared with the current situation, mRNA-1083 was estimated to prevent 2,268 hospitalizations and save €12.1 million in treatment costs. Health and economic benefits were also observed under assumptions of higher COVID-19 rVE, with more than 6,000 hospitalizations prevented and approximately €32 million in treatment cost savings, and under higher COVID-19 incidence, with more than 5,000 hospitalizations prevented and approximately €27 million in treatment cost savings. By contrast, varying influenza incidence, the seasonal influenza CoP-derived rVE, or the egg-adaptation rVE only had a modest impact on model outcomes (**Figure 3**).

**Figure 3.**
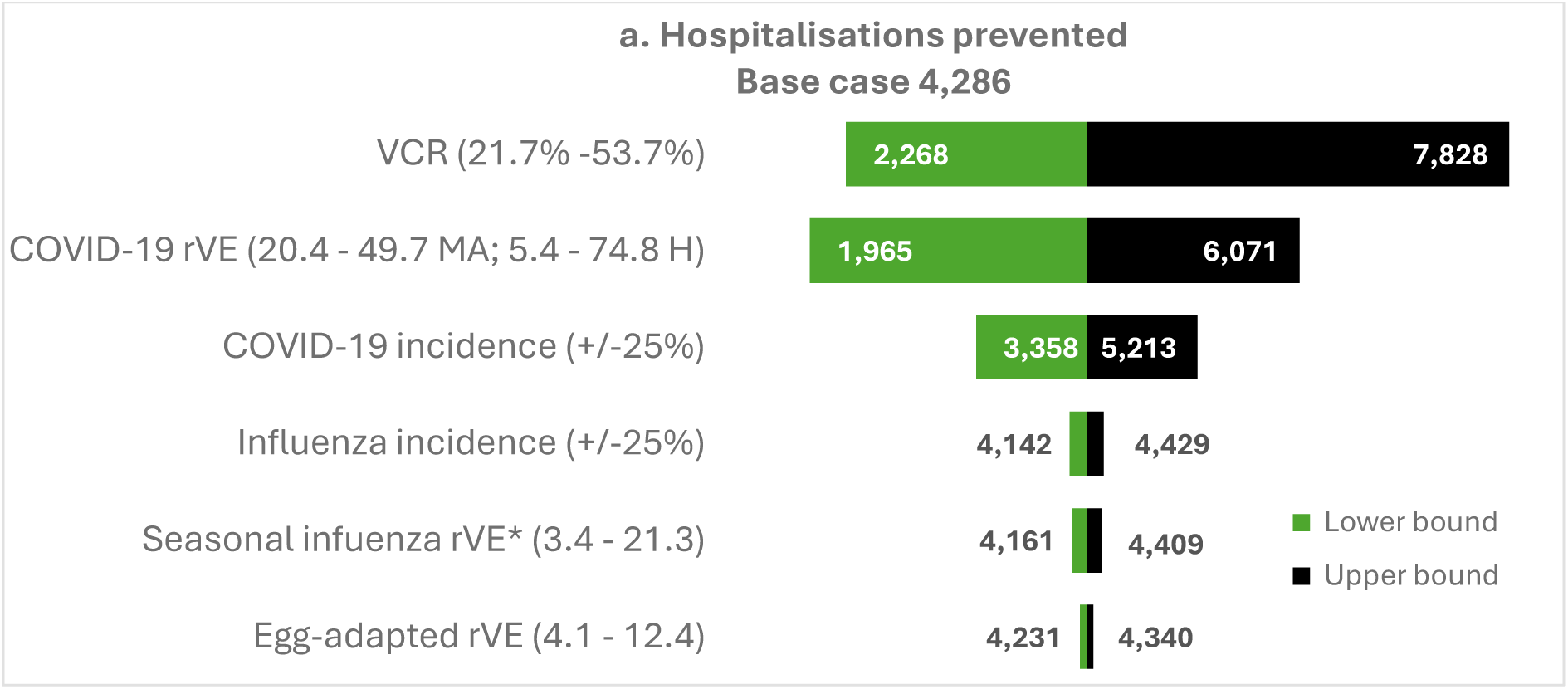
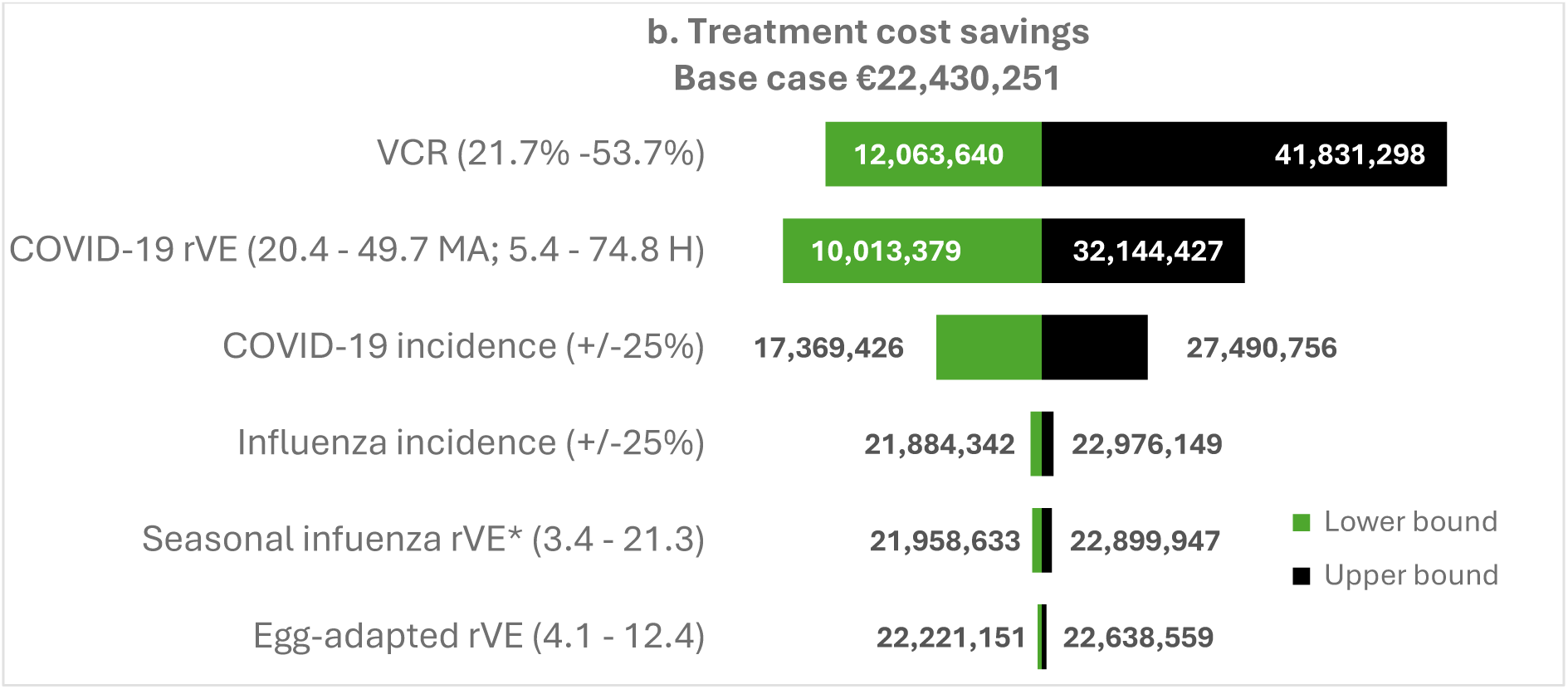
Scenario analysis results for a) hospitalizations prevented and b) treatment cost savings (€) *Seasonal influenza rVE based on the correlate-of-protection; H: hospitalisation; MA: medically-attended; rVE: relative vaccine effectiveness; VCR: vaccination coverage rate

The scenario analysis results for additional outcomes (symptomatic infections, deaths, QALYs, and total healthcare costs) are presented in **Figure S2**. For these additional outcomes, the factors with the largest impact on results were also VCR, COVID-19 rVE, and COVID-19 incidence. The total healthcare cost savings were greater in the low VCR scenario (21.7%) compared with the Step 4 increased VCR (31.7%) (i.e., €73.0M versus €69.4M), as fewer people were vaccinated resulting in fewer vaccination related costs, against moderate cost savings from vaccination driven by lower COVID-19 incidence (**Figure S2**).

## Discussion

Seasonal influenza and COVID-19 continue to place a substantial burden on older adults and the French healthcare system. Despite established vaccination recommendations, coverage against both diseases remains below national targets, leaving a large proportion of the population at risk of severe outcomes. This analysis suggests that introducing the first combination influenza/COVID-19 vaccine, mRNA-1083, could provide meaningful public health and economic benefits by combining operational efficiencies with enhanced clinical protection and increased vaccine uptake.

Compared with separate vaccination using HD-IIV and BNT162b2, mRNA-1083 was estimated to prevent an additional 25,830 medically attended infections, 4,286 hospitalizations, and 1,331 deaths during an average post-pandemic season in France. These health gains translated into annual healthcare cost savings of approximately €69.4 million, comprising €46.9 million in vaccine administration-related costs and €22.4 million in avoided treatment costs. The analysis also demonstrated that the value of mRNA-1083 extends beyond combining two vaccines into a single injection. Potential for increased VE relative to current comparators, the avoidance of egg adaptation, and later influenza strain selection each contributed incrementally to the overall benefits. While the impact of non-egg-based influenza vaccination and better matched strains due to late strain selection was modest in an average season, these advantages may become increasingly important in seasons characterised by egg adaptation or substantial antigenic drift.

The model identified increased vaccination coverage as one of the principal drivers of public health impact. Even under a conservative scenario assuming no improvement in vaccination coverage, mRNA-1083 remained associated with meaningful reductions in hospitalizations and healthcare costs. However, substantially larger benefits were observed when vaccination coverage for COVID-19 increased to match current influenza uptake, highlighting the importance of simplifying seasonal vaccination. The plausibility of increased vaccination coverage with a combination vaccine is supported by preference studies demonstrating that older adults and HCPs favour combination vaccination because of its convenience and reduced number of injections [23].

These findings are particularly relevant in the French context. During the severe 2024/25 influenza season in France, around 100 hospitals activated their *Plan Blanc* emergency response plans because of exceptional pressure on healthcare services, while more than 70% of patients presenting to emergency departments with influenza were eligible for, but had not received, seasonal influenza vaccination [71].

The Haute Autorité de Santé also recommends co-administration of influenza and COVID-19 vaccines to simplify vaccination and improve uptake [17, 72]. A combination vaccine directly supports this strategy by replacing two separate administrations with a single injection, potentially improving the experience for vaccine recipients while reducing HCP workload. A survey among French pharmacists indicate that prescribing and administering a combination vaccine could save approximately 9.5 minutes compared with co-administering separate influenza and COVID-19 vaccines [24, 62]. Our model projected that introducing mRNA-1083 could save approximately 439,000 hours of HCP time each influenza season in France. Such operational efficiencies could be particularly valuable during periods of peak healthcare demand, when workforce capacity is constrained and healthcare systems are under considerable pressure.

The analysis also highlights the potential advantages of the mRNA platform for influenza vaccination. Unlike conventional egg-based vaccines, mRNA vaccines avoid egg-adaptive mutations that may reduce VE, and they can be manufactured more rapidly, enabling later strain selection using more recent surveillance data. Although later strain selection would require changes to the current WHO strain selection process and associated regulatory and policy frameworks, our scenario analysis suggests that implementation of this approach could generate additional public health benefits by reducing the impact of antigenic drift in seasons where vaccine mismatch occurs. Of note, the analysis only considered a crude approximation of these effects by presenting an average across multiple seasons and thus is limited in providing insights into the potential magnitude of effect during a season when egg-adaptive mutations or antigenic drift is occurring.

Several limitations should be considered. First, because influenza and COVID-19 incidence vary considerably between seasons, the model represented an average post-pandemic season using epidemiological data from the two most recent seasons. Second, real-world effectiveness data for mRNA-1083 are not yet available. Vaccine effectiveness was therefore estimated using evidence from pivotal clinical trials, correlate-of-protection analyses for the influenza component [68], and real-world effectiveness estimates for mRNA-1283 against COVID-19 [65]. These estimates should be refined upon availability of mRNA-1083 real-world evidence. Third, utility values were derived from international studies because French-specific estimates were unavailable. The analysis is also likely to underestimate the full value of combination vaccination. The model did not capture the potential benefits of mRNA-1083 on preventing downstream consequences of influenza or COVID-19 infection, including triggering and exacerbating cardiorespiratory disease, long-term functional decline, or loss of independence among older adults [33, 73]. Similarly, indirect societal impacts such as productivity losses among caregivers, disruption of healthcare services during epidemic peaks, or the broader benefits of reducing hospital overcrowding, and ecological impact were not considered [36]. However, the analysis assumed that implementation of a combination vaccine would not affect acceptance among vaccinators, as a reduction in administration fees may lower HCP preference for the combination vaccine.

Overall, this study suggests that mRNA-1083 could substantially reduce the burden of seasonal influenza and COVID-19 in France while generating meaningful healthcare cost savings. By combining potential enhanced protection against both viruses with simplified vaccine delivery, mRNA-1083 has the potential to improve vaccination program efficiency and increase seasonal vaccine uptake. This remains particularly relevant as SARS-CoV-2 has become endemic, with continued high rates of hospitalization and severe outcomes among older adults despite lower incidence than during the pandemic [3, 4]. Increasing vaccination coverage in this population therefore remains a public health priority. Beyond reducing the overall burden of respiratory disease, improving COVID-19 vaccine uptake may also help reduce health inequalities, as social vulnerability has been associated with higher mortality and greater need for mechanical ventilation among patients admitted to intensive care with COVID-19 in France [74]. As the French population continues to age and seasonal respiratory viruses continue to place pressure on healthcare services, combination vaccination with mRNA-1083 represents a promising strategy to strengthen respiratory disease prevention while improving the efficiency and resilience of vaccination programmes.

## Conclusions

mRNA-1083 combines protection against influenza and COVID-19 in a single vaccination and has the potential to deliver substantial public health and economic benefits compared with separate seasonal vaccination. In this analysis, mRNA-1083 was estimated to prevent more than 25,830 medically attended infections and 4,286 hospitalizations, while saving 439,285 HCP hours and generating €69.4 million in healthcare cost savings. By integrating operational efficiencies, enhanced clinical protection, increased vaccination uptake, and the technological advantages of the mRNA platform, mRNA-1083 represents a promising strategy to strengthen seasonal respiratory disease prevention and improve the long-term sustainability and resilience of vaccination programs in France.

## Disclosures

### Author contributions

CD, MU, NvdV, DM, EB, LB, JC and HL were involved in the study design, analysis, writing and critical revisions. External experts (GG, OG, ORL and OR) were involved in the review and critical revisions of the manuscript. All authors read and approved the final manuscript.

### Data availability

All publicly available data generated or analysed during this study are included in this published article and as supplementary information files.

### Medical writing, Editorial, and other Assistance

Medical writing and editorial assistance were provided by Kavi Littlewood (Littlewood Writing Solutions) in accordance with Good Publication Practice (GPP 2022) guidelines, funded by Moderna, Inc., and under the direction of the authors.

### Declaration of generative AI use

The authors report generative AI was not used in their research or preparation of this manuscript.

## Data Availability

All data produced in the present work are contained in the manuscript

