## Supplemental File for "Potential public health and economic impact of influenza and COVID-19 multicomponent vaccination (mRNA-1083) for adults aged 65 years and older in France"

[1]Moderna Inc., Cambridge MA, USA; [a]Affiliation at time of study conduct and manuscript development

### Content:

**Table S1.** Model inputs for the 2023/24 and 2024/25 seasons in adults ≥65 years

**Table S2.** Symptomatic infections, deaths and QALYs prevented in each step

**Figure S1.** Vaccine coverage rates in VCR increase (Step 4) and scenario analyses (COVID-19 VCR and Influenza VCR)

**Figure S2:** Additional scenario analyses results

#### Table S1. Model inputs for the 2023/24 and 2024/25 seasons in adults ≥65 years

| **COVID-19 events** | 2023/24 Season | 2024/25 Season | Median | **Source** |
|---|---|---|---|---|
| Symptomatic infections requiring care | 299,313 | 143,534 | 221,424 | [1-4] |
| Hospitalisations | 36,855 | 17,674 | 27,265 | |
| *With intensive care* | 646 | 326 | 486 | |
| *Without intensive care* | 36,209 | 17,348 | 26,778 | |
| Outpatients | 262,458 | 125,860 | 194,159 | |
| Deaths | 15,923 | 8,234 | 12,078 | |
| *Inpatient* | 2,837 | 1,363 | 2,100 | |
| *Out-of-hospital* | 13,086 | 6,871 | 9,978 | |
| **Influenza events** | 2023/24 Season | 2024/25 Season | Median | [2-4] |
| Symptomatic infections requiring care | 102,481 | 217,935 | 160,208 | |
| Hospitalisations | 7,854 | 17,749 | 12,801 | |
| *With intensive care* | 359 | 851 | 605 | |
| *Without intensive care* | 7,495 | 16,898 | 12,196 | |
| Outpatients | 94,627 | 200,186 | 147,407 | |
| Deaths | 3,286 | 9,092 | 6,189 | |
| *Inpatient* | 558 | 1,268 | 913 | |
| *Out-of-hospital* | 2,728 | 7,824 | 5,276 | |

| **Calibrated model inputs** | Influenza | | COVID-19 | | |
|---|---|---|---|---|---|
| | 65-74y | ≥75y | 65-74y | ≥75y | Calibration |
| Annual incidence symptomatic infections requiring care | 0.70% | 1.72% | 0,74% | 2,49% | |
| Probability of hospitalisation | 7.99%* | | 12,63%* | | |
| Probability of outpatient care | 92.01% | | 87.37% | | |
| Probability of ICU | 6.9% | 3.8% | 2.72% | 1.50% | |
| In-hospital mortality | 3.9% | 8.5% | 4.7% | 8.6% | |
| Out-of-hospital mortality | 1.3% | 4.6% | 3.8% | 5.5% | |
| **Baseline utility** | 65-74 years | | 75 years | | [5] |
| | 0,8100 | | 0,7350 | | |

| **Disutilities** | Average disutility | Duration | |
|---|---|---|---|
| Myocarditis due to COVID-19 | -0.7 | 2 days | [6] |
| Symptomatic influenza infections requiring care | -0.208 | 7 days | [7] |
| Symptomatic COVID-19 infections requiring care | -0.19 | 7 days | [8] |
| Hospitalisation influenza | -0.018 | / | [9] |
| Hospitalisation COVID-19 no ICU | -0.63 | 10.4 days | [10] |
| Hospitalisation COVID-19 with ICU | -0.83 | 18.4 days | [10] |
| Long COVID-19 after outpatient care | -0.278 | 90 days | [11] |
| Long COVID-19 after hospitalisation | -0.278 | 180 days | [11] |

| **Resource use** | | Description | |
|---|---|---|---|
| **Administration cost** | | | |
| Pharmacist fee | €7.50 | Influenza vaccination (including VAT) | [12-14] |
| General practitioner fee | €35.55 | Average cost of Consultation for any act weighted by Health Insurance activity | |
| Nurse fee | €13.45 | Costing: 2.4xAMI+1MAU +1IFD +5.1IK | |
| Average administration cost | €18.83 | Assumes homogeneous distribution of vaccine administration among health professionals | |
| **Outpatient care costs** | | | |
| GP consultation | €35.55 | Average cost of GP Consultation for any act weighted by Health Insurance activity report and including fees | [14] |

| | | | |
|---|---|---|---|
| Paracetamol | €2.09 | | [15] |
| Average outpatient care cost | €37.64 | | |
| **Hospitalisations** | | | |
| Influenza hospitalisation | €3,331.97 | Weighted average price of influenza hospitalisation (PD, valuation from the perspective of AMO, MCO stays) | Scansanté/GHS [16] |
| COVID-19 hospitalisation no ICU | €4,935.23 | Average cost of stay without ICU + medical transport | [17, 18] |
| COVID-19 hospitalisation with ICU | €17,092.70 | Average cost of stay with ICU + medical transport | [17, 18] |

AMI: Acte Médico-Infirmier; AMO: Assurance Maladie Obligatoire; GHS: Groupes homogènes de séjours; GP: general practitioner; ICU: intensive care unit; IFD: Indemnité Forfaitaire de Déplacement; IK: Indemnité Kilométrique; MAU: Majoration d'Acte Unique; MCO: Médecine, Chirurgie, Obstétrique; Primary Diagnosis; VAT: value-added tax

**Table S2. Symptomatic infections, deaths and QALYs prevented in each step**

| | COVID-19 | Influenza | Total |
|---|---|---|---|
| **Symptomatic infections** | | | |
| Step 2 | 10,761 | 1,592 | 12,354 |
| Step 3 | - | 933 | 933 |
| Step 4 | 7,876 | 4,668 | 12,543 |
| Step 5 | - | 1,858 | 1,858 |
| **Deaths** | | | |
| Step 2 | 606 | 62 | 668 |
| Step 3 | - | 36 | 36 |
| Step 4 | 447 | 180 | 627 |
| Step 5 | - | 72 | 72 |
| **Quality-adjusted life-years (COVID-19 and influenza)** | | | |
| Step 2 | 3,891 | | 3,891 |
| Step 3 | 194 | | 4,084 |
| Step 4 | 3,596 | | 7,680 |
| Step 5 | 386 | | 8,066 |

**Figure S1. Vaccine coverage rates in VCR increase (Step 4) and scenario analyses (COVID-19 VCR and Influenza VCR)**

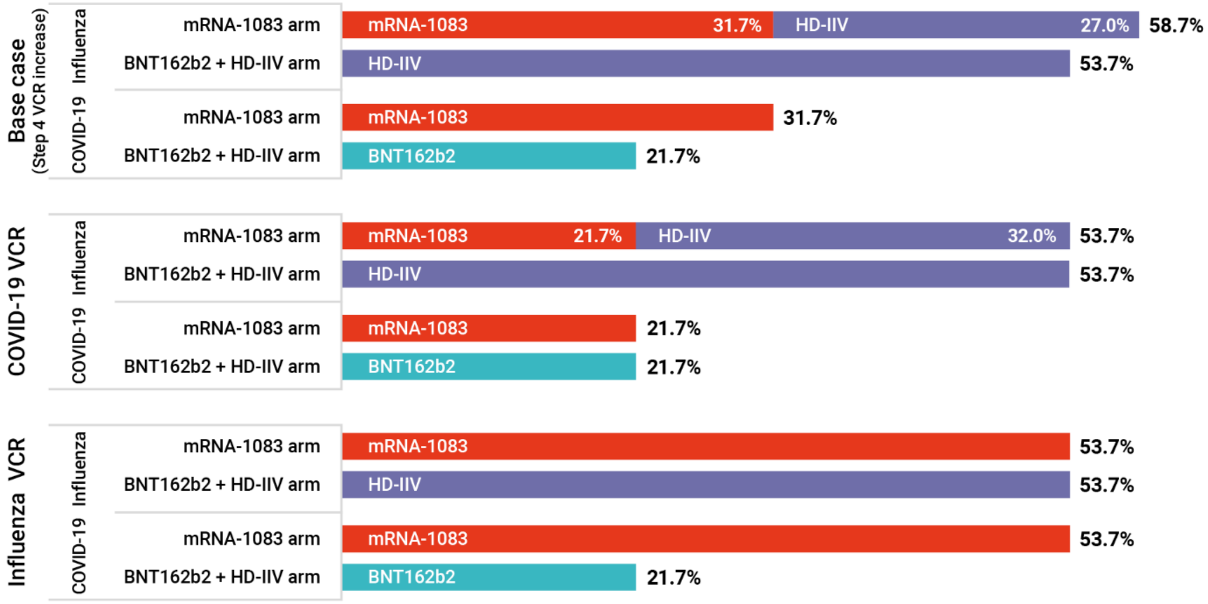


VCR: vaccine coverage rate

**Figure S2. Additional scenario analyses results**

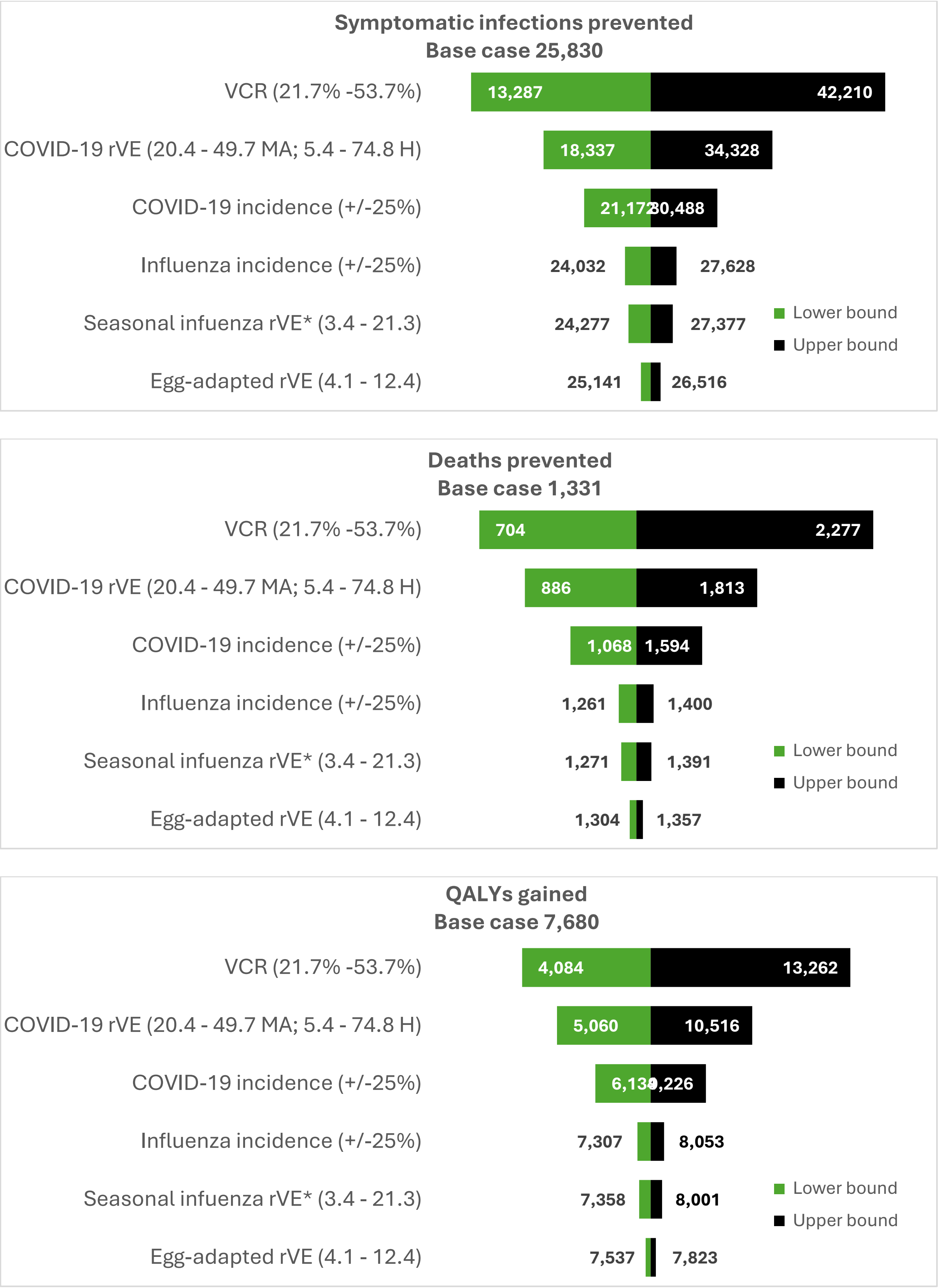

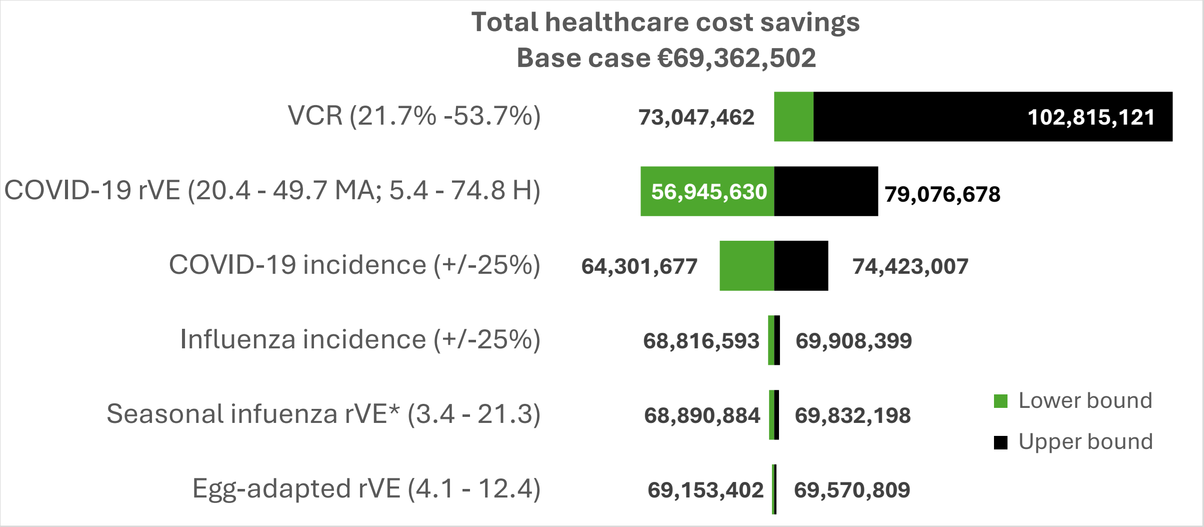


*Seasonal influenza rVE based on the correlate-of-protection; H: hospitalization; MA: medically-attended; rVE: relative vaccine effectiveness; VCR: vaccination coverage rate
